# Generalisability of AI-based scoring systems in the ICU: a systematic review and meta-analysis

**DOI:** 10.1101/2023.10.11.23296733

**Authors:** Patrick Rockenschaub, Ela Marie Akay, Benjamin Gregory Carlisle, Adam Hilbert, Falk Meyer-Eschenbach, Anatol-Fiete Näher, Dietmar Frey, Vince Istvan Madai

## Abstract

**Background:** Machine learning (ML) is increasingly used to predict clinical deterioration in intensive care unit (ICU) patients through scoring systems. Although promising, such algorithms often overfit their training cohort and perform worse at new hospitals. Thus, external validation is a critical – but frequently overlooked – step to establish the reliability of predicted risk scores to translate them into clinical practice. We systematically reviewed how regularly external validation of ML-based risk scores is performed and how their performance changed in external data.

**Methods:** We searched MEDLINE, Web of Science, and arXiv for studies using ML to predict deterioration of ICU patients from routine data. We included primary research published in English before April 2022. We summarised how many studies were externally validated, assessing differences over time, by outcome, and by data source. For validated studies, we evaluated the change in area under the receiver operating characteristic (AUROC) attributable to external validation using linear mixed-effects models.

**Results:** We included 355 studies, of which 39 (11.0%) were externally validated, increasing to 17.9% by 2022. Validated studies made disproportionate use of open-source data, with two well-known US datasets (MIMIC and eICU) accounting for 79.5% of studies. On average, AUROC was reduced by -0.037 (95% CI -0.064 to -0.017) in external data, with >0.05 reduction in 38.6% of studies.

**Discussion:** External validation, although increasing, remains uncommon. Performance was generally lower in external data, questioning the reliability of some recently proposed ML-based scores. Interpretation of the results was challenged by an overreliance on the same few datasets, implicit differences in case mix, and exclusive use of AUROC.

## Introduction

In the intensive care unit (ICU), prognostic scores are used to monitor patients’ severity of illness, predict outcomes, and guide clinical decisions about interventions and resource allocation [1,2]. These scores have quickly become a fixture in modern critical care and have been adopted in hospitals worldwide [3]. Established scoring systems — such as the Acute Physiology and Chronic Health Evaluation (APACHE) [4] or the Sequential Organ Failure Assessment (SOFA) [5] — rely on a small set of carefully selected parameters to identify patients or patient groups at risk of deterioration [6]. This simplicity comes at the cost of crude prognostication and limited accuracy.

The increasing availability of detailed electronic health records (EHR) has opened the door for developing more sophisticated and personalised scores. Machine learning (ML)-based artificial intelligence (AI) has emerged as a promising tool to leverage the wealth of data [7] and ML-based scores have attracted significant interest within the critical care community [8]. A growing body of literature demonstrates improved accuracy in predicting a diverse range of outcomes including all-cause mortality [9,10], sepsis [11,12], kidney injury [13,14], respiratory failure [15], and more [16,17].

Despite their promise, ML-based scoring systems are not without risk. One notable challenge is the potential for “overfitting”, where a system’s performance may become overly reliant on unique characteristics of the original patient cohort used for score development. Such overfitting can lead to inaccurate predictions when the system is used in a new hospital, where the original unique characteristics are no longer present [7]. Thus, external validation on data from previously unseen hospitals is a critical first step in establishing the robustness of these systems and ensuring their reliability across different clinical environments [18,19]. Unfortunately, external validation is often disregarded in practice [8,20], raising concerns about the true potential of ML-based scores in the ICU. Indeed, when a proprietary score for the detection of sepsis was implemented in clinical practice, an independent evaluation showed that it performed much worse than anticipated [21]. There is thus potential for an emerging translational gap, where theoretical benefits and advertised gains are not realised in clinical practice.

This systematic review aims to address this issue by determining how frequently external validation is performed in the literature and whether its use has increased in recent years. We further investigated how the performances of ML-based ICU scoring systems typically changed when applied to data from new hospitals. Our work contributes to the ongoing effort of bringing reliable ML-based scores to the ICU bedside.

## Methods

### Eligibility criteria

Studies were included in the review if they 1) described the development of an ML-based AI model that 2) provided early warning of acute patient deterioration in 3) ICU settings based on 4) structured, routinely collected EHR data. To be included in the meta-analysis of model performance, models further needed to 5) be externally validated on data from a geographically distinct hospital that was not part of the derivation cohort. Following Shillan et al. (2020) [8], ML was defined as “any form of automated statistical analysis or data science methodology”. Clinical events were considered “acute” if they occurred up to 7 days after the time of prediction. A model gave early warning of such an event if the event was not yet known to the treating clinician at the time of prediction. The ICU was defined as “an area with a sole function to provide advanced monitoring or support to single or multiple body systems” [8]. Models could be externally validated as part of the same study that developed the model or in a later publication.

Studies were excluded if they: predicted auxiliary outcomes such as length of stay, risk of readmission, laboratory parameters, or values for imputation; used unsupervised learning methods to identify patient subgroups (unless those subgroups were used as input for supervised prediction); included non-ICU patients without providing separate performance metrics (e.g., by including patients from a general ward); required manual note review or prospective data collection of model features; used medical images or natural language processing of free-text notes; only validated the model on data from hospitals that contributed to the development data (including temporal validation on future data); did not report performance in the development dataset.

### Search strategy

We searched the bibliographic databases Ovid MEDLINE and Web of Science for all full-text, peer-reviewed articles matching our search terms published in the English language before April 29th, 2022. We additionally searched the preprint server arXiv for relevant preprints using a custom computer script (see supplementary material at https://doi.org/10.17605/OSF.IO/F7J46). We included only primary research, excluding reviews and conference abstracts (except for abstracts that were peer-reviewed and paper-length, e.g., from the International Conference on Machine Learning).

We divided our search into three sub-themes: “Machine Learning and Artificial Intelligence”, “Intensive care setting”, and “Patient deterioration”. Articles were considered for screening if they matched all three themes. Notably, no theme was defined for external validation, which was ascertained manually during screening. Details of the search strategy including all search terms can be found in the preregistered study Protocol (www.crd.york.ac.uk/prospero, RecordID: 311514).

In an attempt to identify models that were validated in a separate, subsequent publication, we further performed a reserve citation search using Dimensions AI (https://www.dimensions.ai/), looking for validation papers that referenced a screened record (see supplementary material [22]).

### Study selection

Identified articles were exported from the database as RIS files and imported into the reference management software Zotero (Cooperation for Digital Scholarship; version 6.0.26), where they were deduplicated using Zotero’s semi-automated deduplication tool. Titles and abstracts were independently screened for inclusion by four of the authors (AH, BGC, EMA, PR), with each article being seen by at least two reviewers. For all articles that remained after title and abstract screening, full texts were obtained and independently checked for eligibility by two of the authors (EMA, PR). Before each screening stage, screening was piloted on 25 randomly selected articles. Agreement between authors was assessed using Fleiss’ Kappa [23]. If agreement was found to be unsatisfactory (defined as Kappa < 0.6), decisions were calibrated on another set of 25 articles. If there was uncertainty about the eligibility of an article at any stage of the screening, the article was forwarded to the next stage. Any disagreements were resolved in a consensus meeting. If multiple identified articles describe the same model ̶ e.g., when development and external validation were published in separate articles ̶ the article relating to model validation was included and any missing information on performance in the development dataset was supplemented from the article describing the model development.

### Data collection

Limited data collection was performed for all included studies, covering information on target outcome(s), data sources, and whether or not the study was externally validated. For the subset of externally validated studies, a more detailed data collection was performed in the Numbat Systematic Review Manager [24] using a predefined extraction template (see supplementary material [22]). The template was slightly extended prior to data collection to cover all elements defined in the MINimum Information for Medical AI Reporting (MINIMAR) standard [25]. Data collection was performed independently by two authors (EMA, PR). We extracted the following information for each validated study: target population; inclusion/exclusion criteria; information on the data sources including country of origin, number of hospital sites, cohort size, patient characteristics (age, sex, race, socioeconomic status), outcome prevalence; number and type of input features (e.g., vital signs or laboratory tests); ML algorithm; strategy for dealing with missing data; data splitting; performance metrics and performance in internal and external validation; whether the authors explicitly optimised for across-hospital generalisation; and if the authors provided their computer code with the study (e.g., on GitHub). For studies that reported results for more than one algorithm, the performance of the best algorithm during internal validation was recorded. For studies that reported results for more than one outcome, the performance for both outcomes was recorded if they were sufficiently different (e.g., mortality and sepsis), otherwise the most acute outcome was chosen (e.g., mortality at 24 hours if authors reported both mortality at 24 and 48 hours). If a data item could not be ascertained from the main text or supplementary material of the article, it was recorded as missing and no attempt was made to contact study authors for additional data.

### Statistical analysis

Study characteristics and extracted performance metrics were summarised using descriptive statistics and graphical analysis. Changes over time in the proportion of studies performing external validation were assessed using a Chi-square test for linear trend.

Differences in the area under the receiver-operator characteristic curve (AUROC) were analysed using a random-effects model [26]. Parameters were estimated via a Bayesian linear regression model with a single intercept and a normally distributed random effect per study. We used weakly informative normal priors for the mean and half-Cauchy priors for the scale of the random effects [27]. Due to an observed skewed distribution that might unduly influence the results, the difference was modelled with a Cauchy likelihood, which is less sensitive to outliers [28] and is often used for robust regression [29]. Each study’s sample variance was derived using Hanley’s formula [30]. To explore differences in models estimating mortality — which is a well-defined and well-captured ground truth compared to inferred complications such as sepsis [31] or kidney injury [32] — a second model with a fixed effect for mortality was specified. After estimation, we further calculated the proportion of studies in which the absolute difference in AUROC was > ±0.05. A 0.05 threshold was chosen in line with previous studies [33]. No analysis of heterogeneity between studies or risk of bias was performed.

All analysis was performed in R version 4.2.2 [34]. Bayesian linear models were fitted with Hamiltonian Monte Carlo (HMC) using the rstan package version 2.21.8 [35]. All results from the database search, screening, full-text review, and data collection as well as the analysis code are available at the Open Science Framework [22]. A study protocol was pre-registered on PROSPERO (www.crd.york.ac.uk/prospero, RecordID: 311514).

## Results

We identified 4,677 records from MEDLINE (2,613 records), Web of Science (1,863 records), and arXiv (201 records). A detailed flow diagram is shown in Figure 1. After deduplication, the titles and abstracts of 3,851 records were screened. Full texts were assessed for 527 manuscripts, of which 355 (67.4%) described the prediction of acute deterioration in adult ICU patients from routine data (*included studies*). The main reasons for exclusion were prospective or other non-routine data capture, non-acute outcomes, or the inclusion of image, text, or waveform data (Figure 1). Of all included studies, 39 (11.0%) were also externally validated (*validated studies;* Table 1). No additional validation studies were identified through the reverse citation search. Agreement between reviewers as measured by Fleiss’ Kappa was 0.623 for screening and 0.725 for full-text review.

**Figure 1.**
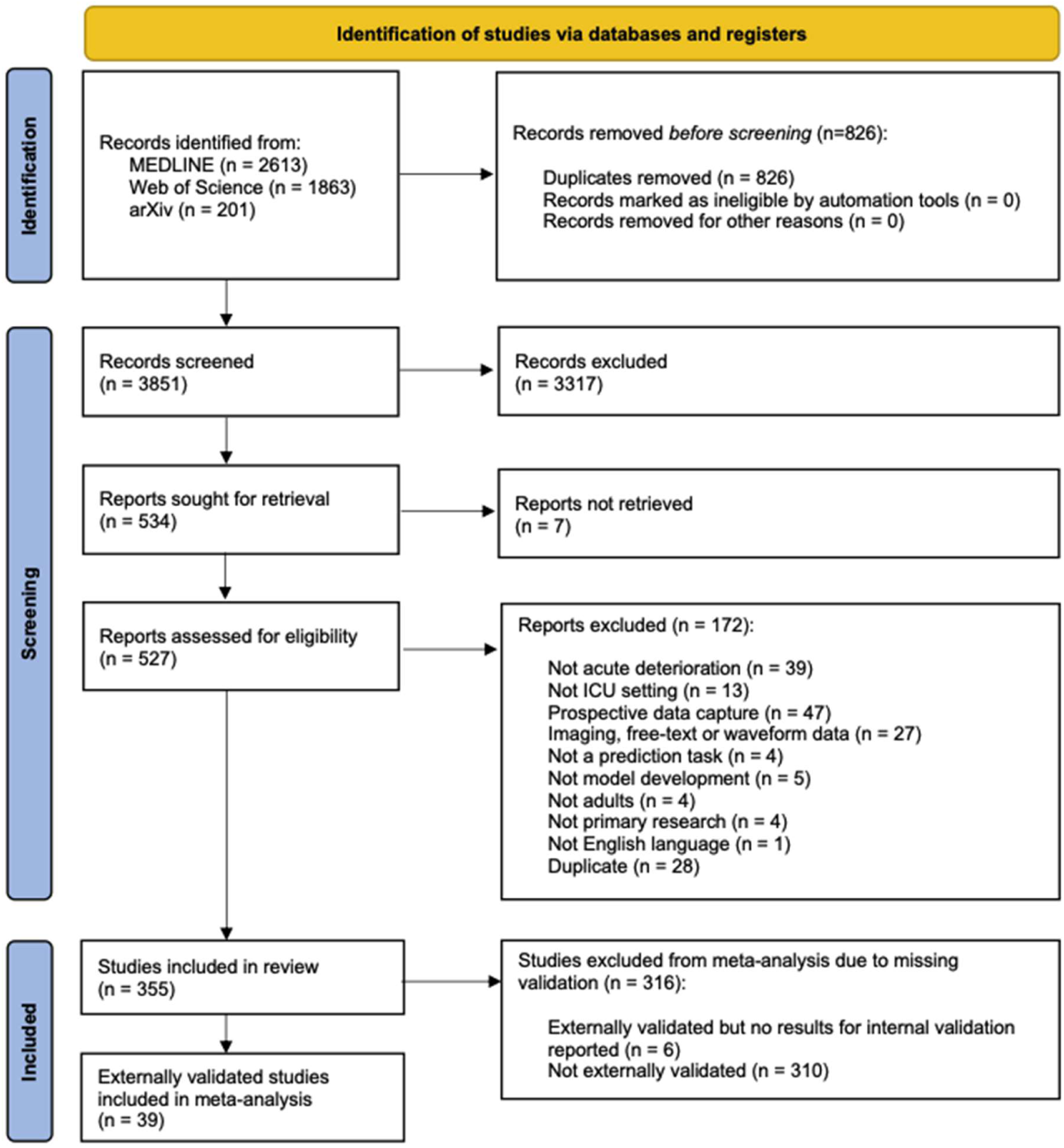
PRISMA flow diagram

**Table 1.** Summary of externally validated studies.

| Authors | Year | Cohort | Outcome | Data source |  | Sample size |  | AUROC |  |
| --- | --- | --- | --- | --- | --- | --- | --- | --- | --- |
|  |  |  |  | Dev | Val | Dev | Val | Dev | Val |
| Pirracchio [9] | 2015 | All patients | Mortality | MIMIC | Other (France) | 25k | 200 | .88 | .94 |
| Delahanty [48] | 2018 | All patients* | Mortality | Other (US) | Other (US) | 147k | 90k | .95 | .94 |
| Moon [49] | 2018 | All patients* | Neurological | Other (Korea) | Other (Korea) | 3k | 325 | .90 | .72 |
| Huang [50] | 2019 | All patients* | Mortality | eICU | +,‡ | 28k |  | .74 | .68 |
| Liu [51] | 2019 | Sepsis | Sepsis (shock) | MIMIC | eICU | 15k | ? | .93 | .85 |
| Shickel [52] | 2019 | All patients* | Mortality | Other (US)<br>MIMIC | + | 36k<br>49k | + | .91<br>.91 | .90<br>.90 |
| van Wyk [53] | 2019 | All patients* | Sepsis | Other (US) | + | 586 | + | - | - |
| Nielsen [54] | 2019 | All patients* | Mortality | Other (DK) | Other (DK) | 10k | 2k | .79 | .73 |
| Kang [55] | 2020 | All patients* | Mortality | MIMIC<br>eICU | + | 21k<br>198k | + | .90<br>.87 | .86<br>.72 |
| Zhao [56] | 2020 | Sepsis | Other | MIMIC | eICU | 11k | 35k | .87 | .84 |
| Roimi [57] | 2020 | Susp.<br>bacteraemia | Infection | MIMIC<br>Other (Israel) | + | 2k<br>1k | + | .89<br>.92 | .59<br>.60 |
| Hyland [16] | 2020 | All patients* | Circulatory | HiRID | MIMIC | 36k | 9k | .94 | .90 |
| Reyna [43] | 2020 | All patients* | Sepsis | MIMIC<br>Other (US) | Other (US) | 20k<br>20k | ? | .82<br>.86 | .81 |
| Wang [58] | 2020 | All patients* | Renal | Other (China) | MIMIC | 11k | 46k | .81 | .95 |
| Liu [59] | 2020 | MODS | Mortality | MIMIC<br>eICU | Other (China) | 15k<br>34k | 439 | .86<br>.85 | .84 |
| Zhou [60] | 2020 | Viral pneumonia | Mortality | eICU | MIMIC | 4k | 937 | .77 | .66 |
| Rahman [61] | 2021 | All patients* | Circulatory | eICU | MIMIC | 216k | 16k | .82 | .90 |
| Zhi [62] | 2021 | Sepsis | Mortality | MIMIC | Other (China) | 2k | 125 | .75 | .54 |
| Holder[63] | 2021 | Sepsis | Other | Other (US) | Other (US) | 9k | 5k | .81 | .77 |
| Hur [17] | 2021 | All patients* | Neurological | Other (Korea) | MIMIC | 12k | 2k | .92 | .70 |
| Chen [64] | 2021 | All patients* | Renal | MIMIC | Other (China) | 46k | 226 | .83 | .79 |
| He [65] | 2021 | Sepsis and AKI | Renal | Other (China) | MIMIC | 209 | 509 | 1.0 | 1.0 |
| Shashikumar [66] | 2021 | All patients* | Sepsis | Other (US) | Other (US) | 17k | 46k | .95 | .93 |
| Levi [67] | 2021 | GI bleeding | Other | MIMIC<br>eICU | + | 4k<br>10k | + | .81<br>.79 | .76<br>.80 |
| Ding [68] | 2021 | Sepsis and AKI | Renal | MIMIC | eICU | 7k | 3k | .70 | .70 |
| Huang [69] | 2021 | AKI | Mortality | MIMIC | eICU | 4k | 1k | .91 | .82 |
| Singhal [70] | 2021 | COVID-19 | Respiratory | Other (US) | Other (US) | 6k | 611<br>77 | .90 | .85<br>.88 |
| Sung [71] | 2021 | All patients* | Mortality | Other (Korea) | Other (Korea) | 22k | 2k | .99<br>.77<br>.84 | .96<br>.77<br>.80 |
| Liu [72] | 2021 | All patients* | Sepsis | Other (US) | eICU | 882 | 6k | .72 | .78 |
| Shawwa [73] | 2021 | All patients* | Renal | Other (US) | MIMIC | 98k | 19k | .69 | .66 |
| Mamandipoor [74] | 2021 | All patients* | Other | eICU | MIMIC | 17k | 13k | .84 | .83 |
| Moor [45] | 2021 | All patients* | Sepsis | MIMIC<br>eICU<br>HiRID<br>AUMCdb | + | 37k<br>57k<br>27k<br>16k | + | .83<br>.80<br>.83<br>.92 | .71<br>.75<br>.73<br>.81 |
| Peng [75] | 2022 | CHF | Renal | MIMIC | eICU | 9k | 10k | .81 | .82 |
| Luo [76] | 2022 | CHF | Mortality | MIMIC | eICU | 6k | 1k | .83 | .81 |
| Kim [77] | 2022 | Cardiac arrest<br>and MV | Mortality | eICU | MIMIC | 2k | 86 | .83 | .76 |
| Fu [78] | 2022 | Cardiogenic<br>shock | Renal | MIMIC | eICU | 1k | 1k | .82 | .73 |
| Zhang [79] | 2022 | Cerebrovascular<br>disease | Renal | MIMIC | Other (China) | 3k | 499 | .88 | .78 |
| Jiang [80] | 2022 | Sepsis | Other | Other (China) | MIMIC | 1k | 688 | .89 | .77 |
| Liang [81] | 2022 | All patients* | Renal | Other (China)<br>MIMIC | AUMCdb | 6k<br>37k | 15k | .86<br>.86 | .87 |

\* "All Patients" are defined as a general adult ICU patient population without specifying additional health conditions (e.g., admitted with sepsis). + Datasets were used alternately for development and validation. ‡ eICU includes data from 208 different hospitals and may thus be used for both development and validation if split by hospital.
AKI, acute kidney injury; AUROC, area under the receiver operating characteristic; CHF, congestive heart failure; DK, Denmark; GI, gastro-intestinal; MODS, Multi-organ dysfunction syndrome; MV, mechanical ventilation; US, United States

### Trend over time

The number of both included and validated studies increased significantly over time (p=0.014) and especially after 2018, with 302 / 355 (85.1%) respectively 38 / 39 (97.4%) studies published in or after that year (Figure 2). The earliest study performing external validation was published in 2015. Between 2018 and 2022, the proportion of validated studies increased from 2 / 28 (7.1%) to 7 / 32 (17.9%; only counting studies published until April 2022).

**Figure 2.**
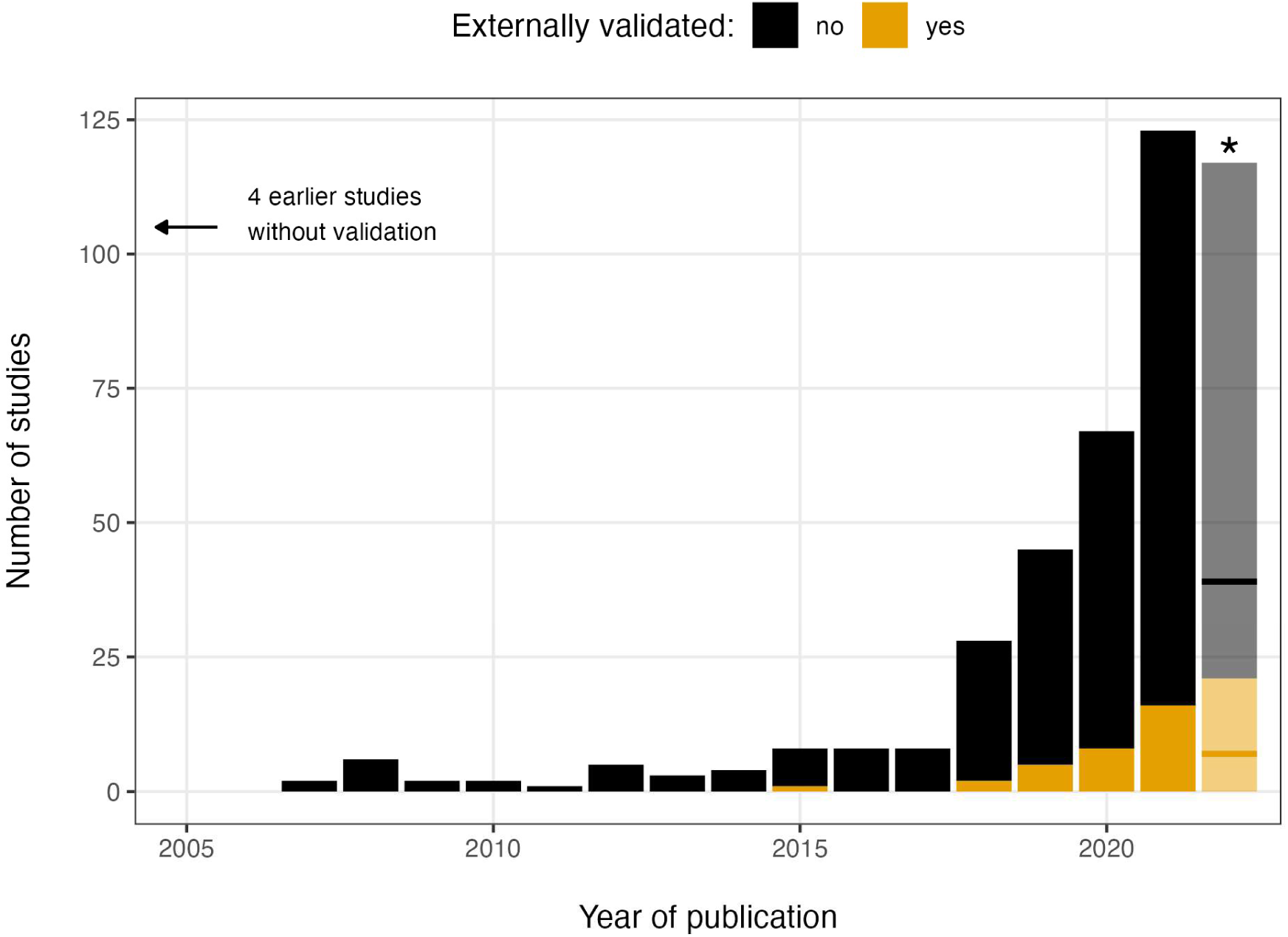
Number of eligible (black) and included externally validated studies (orange) by year of publication. * For the year 2022, only studies published before May were included. Horizontal lines represent the numbers observed until the end of April, with the transparent bar representing the projected numbers for the full year.

### Outcomes

A total of 214 / 355 (60.3%) included studies predicted short-term mortality. The next most commonly predicted outcome was sepsis with 53 / 355 (14.9%), followed by 37 / 355 (10.4%) studies predicting renal complications including acute kidney injury, 19 / 355 (5.4%) studies predicting respiratory complications, 16 / 355 (4.5%) studies predicting circulatory failure, and 14 / 355 (3.9%) studies predicting cardiovascular complications. At only 14 / 214 (6.5%), the rate of external validation was notably lower among studies predicting mortality compared to all studies. If studies predicting mortality were excluded, the proportion of studies that were externally validated — and therefore included in the meta-analysis — almost doubled from 11.0% (39 / 355) to 18.2% (29 / 159).

### Sources of data

Externally validated studies overwhelmingly used US data, with 37 / 39 (94.9%) including studies using at least one US dataset for model development or external validation. Eight studies used Chinese data, 5 studies used European data (Netherlands, Switzerland, Denmark, France), 3 used South Korean data, and 1 used Israeli data.

The publicly available datasets MIMIC [36] and eICU [37] were overrepresented among validated studies. MIMIC was used in 29 / 39 (74.4%) of validated studies compared to 206 / 355 (58.0%) of all included studies, with 14 studies using it for model development, 10 for external validation, and 5 for both. eICU was used in 17 / 39 (43.6%) of externally validated studies compared to 57 / 355 (16.1%) of all included studies, 5 times for model development, 8 times for external validation, and 4 times in both capacities. Together, MIMIC and eICU were used in 31 / 39 (79.5%) validated studies, of which they were the only source of data in 12 / 39 (30.8%) studies. AUMCdb [38] and HiRID [16] — two further, more recent public ICU databases — were only used in 2 / 39 (5.1%) included studies each.

### Performance at new hospitals

All but one of the 39 validated studies reported AUROC. After accounting for sampling variability, model performance in the external validation data was -0.037 (95% credible interval [CI] -0.064 to -0.017; p-value < 0.001) lower than estimated in the internal validation data (Figure 3). Changes in performance ranged from a maximum increase of 0.14 to a decrease of -0.32. In 38.6% of cases, performance loss was < -0.05. On the other end of the spectrum, performance *increased* by > 0.05 in 9.1% of cases – indicating differences in patient populations between train and evaluation cohorts. There was no evidence for differences between studies predicting death and those that predicted other outcomes (p-value = 0.742).

**Figure 3.**
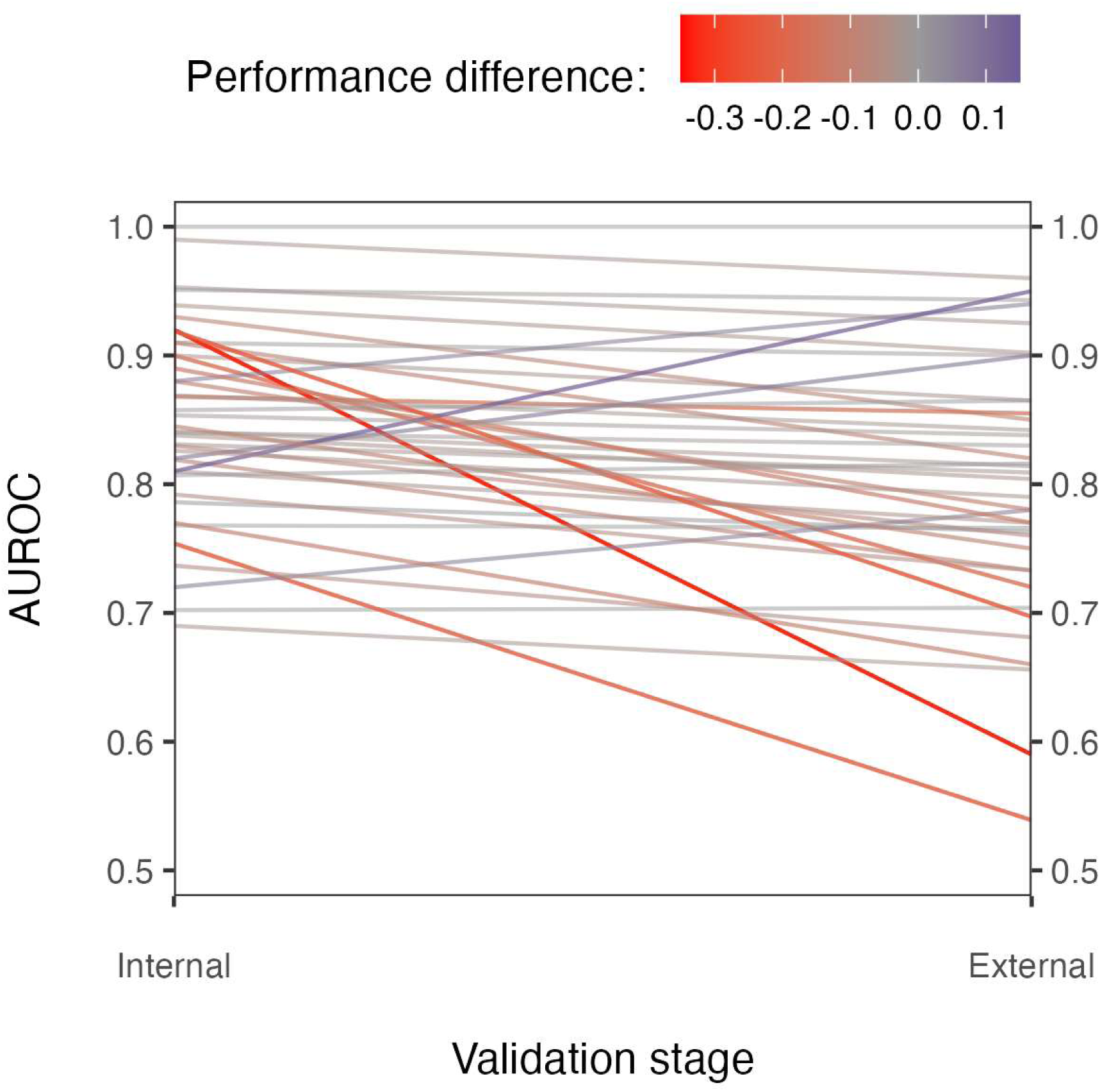
Reported AUROCs for internal and external validation among (N = 39 -1) included studies. One study was omitted because they did not report AUROC.

Other commonly reported metrics included specificity (18 / 39; 46.2%), sensitivity (17 / 39; 43.6%), positive predictive value (16 / 39; 41.0%), F1 score (11 / 39; 28.2%), and accuracy (10 / 39; 25.6%), although they were reported at a much lower rate than AUROC.

## Discussion

This systematic review examined the generalisation of complex, ML-based ICU scoring systems to new hospitals. We considered any score that supports ICU staff through the prediction of imminent patient deterioration from routinely collected EHR data. Leveraging EHR data in this way to improve critical care continues to attract significant research interest, as evidenced by a steady increase in research output. Yet, translating this research into widespread clinical practice — and eventually converting it into patient benefit — requires comprehensive validation of findings, including an evaluation of the scores’ performance at new hospitals. We found that such external validation is still relatively uncommon. Where validation was performed, performance at the new hospital tended to be lower than in the training cohort, often notably so.

### Implications for the translation of AI into clinical practice

Fueled by recent advances in natural language processing and their successful translation to consumer products, there is a reinvigorated hype around the implementation of AI in healthcare [39]. Yet, while many preliminary results keep making the headlines, the proof is in the pudding: a large majority of published results are exploratory in nature, providing only proof-of-concepts [40]. There is a continued lack of verification and clinical validation, blocking the translation of these proof-of-concepts to actual products [19]. In our review, we demonstrate that the issue of inadequate verification extends to ML-based scoring systems: the rate of retrospective external validation – a first step to establish validity and robustness – remains low. Less than 20% of identified studies that proposed new scoring systems for the ICU underwent external validation. External validation in this context is an essential step for clinical adoption. Unless a model is solely built for use in the hospital(s) it was developed at – an unlikely goal – it should be judged by its accuracy across a range of hospitals, all of which may potentially use the model in the future. When evaluated this way, we found that average model accuracy as measured by the AUROC reduced by -0.037 compared to the training hospitals. This constitutes a relative decrease of 7-23% in performance, with decreases of up to and more than 50% in some cases. Many ostensibly well-performing scores may thus no longer be suitable for use at the new hospital, a fact that would (and does) go unnoticed in the absence of external validation. To actually facilitate translation to the clinical setting, rigorous external validation must become the standard when developing ML-based scoring systems and clinical AI more generally. Retrospective external validations in particular aid the early identification of model deficiencies, highlighting the need for training and fine-tuning on a broader variety of training data [41]. While there is still a long way to go to make such external validation the default, our review at least suggests that there is a growing recognition of its importance among researchers: over 80% of all identified studies performing external validation were published in 2018 or later.

### Interpretation of external validation results

The infrequent external validation of ML models for the prediction of acute events in the ICU was already noted in a 2019 systematic review, with only 7% of studies at the time using geographically independent data for model validation [8]. This has been echoed in more recent, disease-specific reviews looking at models for sepsis [20] and acute kidney injury [42]. While we showed that this percentage has somewhat improved since, we also find that challenges remain even if external validation is performed.

While we observed a tendency for reduced model performance in external data, the magnitude of reduction was milder than anticipated from previous studies [41,43–45]. This may partially be explained by the performance metric. We focused on the AUROC as the primary effect measure, since it allowed performing a meta-analysis due to its popularity and its comparability across different levels of prevalence. However, AUROC may be less sensitive to changes in the data. For example, while the drop in AUROC in the PhysioNet CinC challenge 2019 [43] was generally mild and in line with our findings, the “utility of prediction” — a custom metric defined as a timely prediction within 12 hours before to 3 hours after the onset of sepsis — in the new hospital was worse than not predicting at all.

The average reduction in performance might have been more pronounced if another metric such as utility or normalised AUPRC were used instead of AUROC. Unfortunately, it was not possible to include such metrics in a meta-analysis due to their infrequent reporting. We recommend that future validation studies systematically report multiple performance metrics that represent the performance holistically.

The observed moderate reduction in average performance may have also been driven by the non-negligible number of models whose performance *increased* during external validation. Whereas minor fluctuations may occur due to sampling variability, a model’s performance shouldn’t notably increase in the external validation cohort. If it does, this suggests that there may be systematic differences in case mix between the training and validation cohorts – rendering the performances incomparable. If cohorts cannot be defined well enough to ensure their comparability, we recommend also reporting the performance of a model trained solely on the validation data. This provides a (potentially overfit) upper limit on what might have been achieved in the external data [41] and thus allows readers to take any distorting effects of case mix into consideration.

Finally, although the rate of external validation is slowly rising, it appears almost exclusively confined to a few open-source validation sets, most prominently MIMIC [36] and eICU [37]. A version of MIMIC was used in almost 80% of all identified studies that performed external validation. This is potentially problematic, as studies worldwide are thus largely judged by their ability to retain performance in patients from the single US hospital included in MIMIC, which does not necessarily represent the wider ICU population. This means that users and reviewers need to closely scrutinise claims of external validation in the area of ICU scoring systems if they judge tools that are to be used outside of the specific clinical settings captured by MIMIC. This also highlights that while large open-source datasets are able to fuel a large number of publications in certain areas, they do not necessarily by themselves improve the ability to build models that generalize, limiting their impact on successful translation to the clinical setting.

### Strengths and limitations

We used a thorough, pre-defined search strategy to identify all relevant studies, covering two major bibliographic databases as well as the most relevant preprint server for ML research. Inclusion criteria were carefully assessed for all identified records by at least two reviewers and we additionally performed a reverse reference search to ensure we did not miss validation results that were published as stand-alone manuscripts.

To allow for direct comparability of AUROC in the development and validation data, we limited our analysis to external validation on retrospective, routine data. We did not capture validation that was performed by prospectively collecting additional data or within clinical trials. This has two important implications. First, the proportion of validated studies may be higher than reported here, especially in the years preceding the availability of large open-source datasets. Second, the reported performances do not imply clinical usability but rather reflect the stability of study results across different sets of data. Nevertheless, external validation in retrospective data is an invaluable first step to assess the usability of a prediction model in clinical practice and should be considered for any study developing prediction models from routine data. Existing findings are fundamental to the conception of future studies and basing future research on ‘false’ or non-robust results can significantly hinder genuine innovation in the field, creating a substantial drain on both time and financial resources.

Due to the anticipated heterogeneity of studies, we limited ourselves to a descriptive summary of study results and trends. We did not perform a risk of bias assessment. Previous studies that assessed study quality reported a neglect of model calibration, inappropriate internal validation, and overall lack of reproducibility [20,42], all of which may also have been presented in the studies included here. Our results also assume that there were no systematic differences between studies that did and did not get externally validated. This is a strong assumption. For example, studies that were externally validated may be more generalisable to begin with because good performance in new dataset(s) was an explicit part of the study objectives. In this case, the true performance drop among non-validated studies may be even greater than estimated here.

## Conclusion

Given the increasing availability of routine data capture, open-source ICU data sources [16,36–38], and well-documented tools for data harmonisation and preprocessing [46,47], there is little standing in the way of external validation of ML-based scoring systems. External validation should thus become the default for any study developing new scoring systems. It can provide invaluable information on the robustness of newly proposed scores and their potential for widespread adoption. However, while some external validation is certainly better than none, any results derived from it will only truly be useful if the data used for validation is representative of the model’s intended future use setting. The data used for validation should be carefully selected and interpreted to ensure a fair comparison and enable meaningful interpretation, taking into account shifts in data quality, patient case mix, and any other factors that may impact model performance.

## Data Availability

All data produced are available online at https://doi.org/10.17605/OSF.IO/F7J46.

